# Early Pandemic Associations of Latitude, Sunshine Duration, and Vitamin D Status with COVID-19 Incidence and Fatalities: A Global Analysis of 187 Countries

**DOI:** 10.1101/2024.11.29.24318208

**Authors:** Reagan M. Mogire

**Affiliations:** Center for Research On Genomics and Global Health, National Human Genome Research Institute, National Institutes of Health, Bethesda, MD, USA; Center for Clinical Research, Kenya Medical Research Institute (KEMRI), P. O. Box 54, Kisumu, 40100, Kenya

**Keywords:** COVID-19, latitude, sunshine, vitamin D, prevalence, mortality rate, case fatality rate, early pandemic, intervention measures

## Abstract

In the face of the COVID-19 pandemic, understanding the interplay between environmental factors and virus spread is crucial for global preparedness strategies. This study explores how geographic latitude, sunshine duration, and vitamin D status were associated with the incidence and fatality rates of COVID-19 across 187 countries during the crucial early months of the outbreak. Data on the total number of COVID-19 cases by country were obtained from the United Nations database as of June 30, 2020. Univariate and multivariate regression analyses were conducted to determine the associations between COVID-19 cases and latitude, average hours of sunshine from January to June, and mean 25-hydroxyvitamin D (25(OH)D) levels. The average COVID-19 prevalence and mortality per million population were 2,087 and 69, respectively, with a case fatality rate of 3.19%. COVID-19 case fatality rate was positively associated with latitude (β = 0.030; 95% CI: 0.008, 0.052) and negatively associated with hours of sunshine (β = -1.51; 95% CI: -4.44, 1.41) and 25(OH)D levels (β = - 0.054; 95% CI: -0.089, -0.019) in adjusted regression analyses. Findings were similar for COVID-19 prevalence and mortality rate. These findings indicate that higher latitude and lower 25(OH)D levels was associated with increased COVID-19 severity and mortality.

While the data highlight potential links between vitamin D status and COVID-19 outcomes, causality cannot be inferred. Further research, including large-scale, well-controlled trials, is essential to determine whether vitamin D plays a definitive role in COVID-19 prevention and management.

## Introduction

As of 2024, the COVID-19 pandemic caused by the novel coronavirus SARS-CoV-2 continues to pose significant global health challenges^1,2^. Despite advancements in vaccination and treatment strategies, the virus remains endemic in many regions due to factors such as viral mutations, vaccine hesitancy, and unequal access to healthcare resources^3,4^. The persistent circulation of COVID-19, along with the possibility of future SARS-like epidemics, underscores the urgent need to understand the factors influencing the incidence, transmission, and severity of such infectious diseases^5^.

COVID-19 has resulted in substantial morbidity and mortality worldwide. By October 2024, the World Health Organization reported over 770 million confirmed cases and nearly 7 million deaths globally^1^. While factors like age, sex, and underlying comorbidities (e.g., hypertension, diabetes, obesity) have been identified as key determinants of disease severity^6^, disparities in COVID-19 incidence and outcomes among different countries and populations suggest that additional factors may contribute to these variations^7^.

Notably, some countries with limited healthcare infrastructure and high population densities in Africa experienced relatively lower COVID-19 incidence and mortality rates than other countries ^8^. In contrast, populations of African descent residing in temperate regions such as the United States and the United Kingdom have been disproportionately affected, exhibiting higher rates of severe disease and mortality compared to other ethnic groups^9,10^. These disparities have prompted investigations into environmental and nutritional factors, including the potential role of vitamin D status, that may influence susceptibility to and outcomes from COVID-19^11,12^.

Vitamin D deficiency has emerged as a potential modifiable risk factor affecting COVID-19 outcomes^13,14^. Vitamin D, primarily obtained through skin synthesis upon exposure to ultraviolet B (UVB) radiation from sunlight, plays a crucial role in modulating immune function^15^. Several studies have suggested that low vitamin D levels may be associated with an increased risk of respiratory infections, including COVID-19^16,17^. Moreover, geographical factors such as latitude and sunshine duration, which influence the endogenous production of vitamin D, may contribute to the observed differences in COVID-19 incidence and mortality rates across regions^18^.

Previous research examining the associations between COVID-19 outcomes and factors like latitude, sunshine exposure, and vitamin D status has often been limited to specific cities or regions, potentially restricting the generalizability of the findings^19,20^. To address this gap, a comprehensive analysis was conducted on data from 187 countries to investigate the relationships between COVID-19 prevalence, mortality rate, case fatality rate (CFR), and factors such as latitude, sunshine duration, and mean vitamin D status. Understanding these associations is critical not only for enhancing current public health interventions against COVID-19 but also for preparing for future respiratory epidemics^21,22^. Insights gained from this study may inform strategies for prevention and management, including nutritional interventions and policies aimed at mitigating the impact of COVID-19 and similar infectious diseases.

## Methods

This study utilized open-access COVID-19 data from the COVID-19 Data Repository by the Center for Systems Science and Engineering (CSSE) at Johns Hopkins University, available at https://github.com/CSSEGISandData/COVID-19. The dataset includes daily cumulative cases and deaths from January 2020, when only China had reported cases, up to June 30, 2020, covering the end of winter in the Northern Hemisphere on June 21, 2020.

COVID-19 data were primarily analyzed as of June 30, 2020, serving as the central reference point for this study. For comparative analysis, data from two additional dates—March 31 and September 30, 2020—were also examined. Multiple entries for each country, including data reported for individual states or provinces, were aggregated to provide a single, comprehensive value for each country. Country population data were obtained from the United Nations’ World Population Prospects 2019 Revision^23^. Monthly average sunshine duration data for each country were sourced from the World Meteorological Organization (WMO)^24^. Mean 25-hydroxyvitamin D (25OHD) levels for countries were gathered from existing literature, prioritizing national surveys, followed by community-based studies or country-level data in that order.

For the analyses, prevalence was defined as the number of COVID-19 cases per one million population in a country. The mortality rate was defined as the number of deaths per one million population. The case fatality rate (CFR) was calculated as the percentage of confirmed COVID-19 patients who died (number of deaths divided by the number of confirmed cases, multiplied by 100) in a country.

Statistical analyses were conducted using R version 4.0.2 (R Foundation for Statistical Computing, Vienna, Austria), available at https://www.R-project.org/. Variable normality was evaluated using the Kolmogorov-Smirnov test. To enable parametric analyses, numbers of COVID-19 cases and deaths, along with other parameters, were log-transformed to achieve normal distribution. However, mean values were calculated from untransformed data for descriptive statistics. Associations between COVID-19 metrics—prevalence, mortality rate, and case fatality rate—and factors such as latitude, average sunshine duration, and mean 25-hydroxyvitamin D (25OHD) levels were explored using univariate and multivariate linear regression models. The multivariate models were adjusted for gross domestic product (GDP—to control for socioeconomic factors that might influence the COVID-19 outcomes and testing), elderly dependency ratio (EDR—to account for the higher impact of COVID-19 on elderly populations), and population density (to adjust for the increased transmission rates in denser populations), with data sourced from the relevant databases. Scatter plots visualized these relationships for each country, with a p-value of less than 0.05 denoting statistical significance. For analyses involving geographic position, latitudes north of the equator were coded as positive values, indicating greater distance from the equator, while southern latitudes were coded as negative, indicating proximity to the equator. This coding reflects the seasonal variation in sunlight exposure relevant until June, after which the seasons significantly change.

## Results

### Population Characteristics

This study analyzed data up to June 30^th^ from 187 countries across the seven UN world regions (Figure 1 and Supplementary Figure 1). The dataset included 10,446,927 confirmed COVID-19 cases and 508, 459 deaths. The average COVID-19 prevalence and mortality rates per million population were 2,087 and 69, respectively, with an overall CFR of 3.19%.

**Figure 1.**
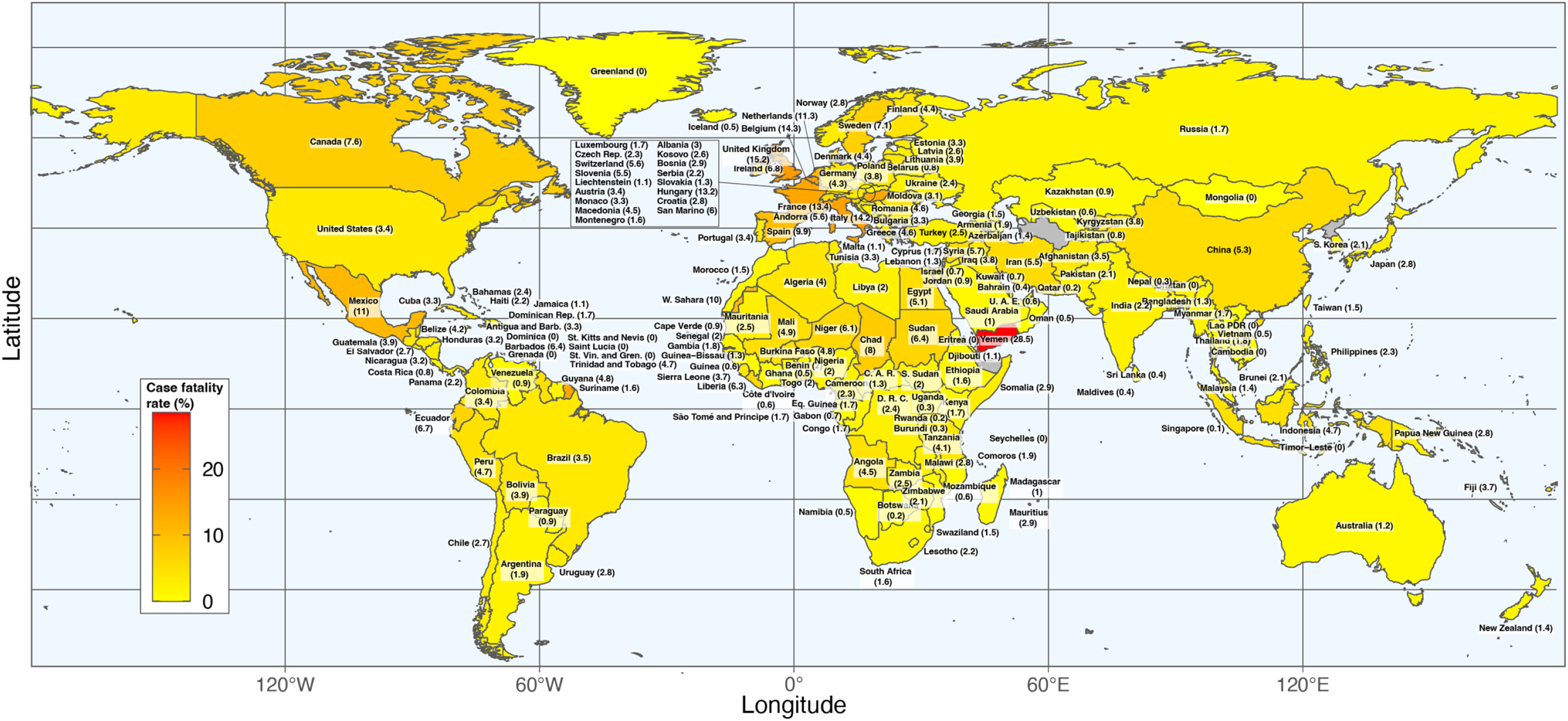
Case fatality rate (%) of COVID-19 by country as at 30^th^ June 2020.

### Effect of latitude on COVID-19 prevalence, mortality rate, and case fatality rate

Linear multivariate regression analyses demonstrated a positive correlation between latitude and COVID-19 prevalence (β = 0.012; 95% CI: 0.001, 0.024), mortality rate (β = 0.042; 95% CI: 0.014, 0.070), and CFR (β = 0.030; 95% CI: 0.013, 0.050) (Table 2). A similar trend was observed up to the end of March 2020 but not significant in September 2020 (Supplementary Table 1). Scatter plots also showed a positive correlation between latitude and COVID-19 metrics (Figure 2A, B and C). Latitude accounted for 5%, 16%, and 4% of the variation in COVID-19 prevalence, mortality rate, and CFR, respectively by the end of June 2020 (Figure 2).

**Table 1.**
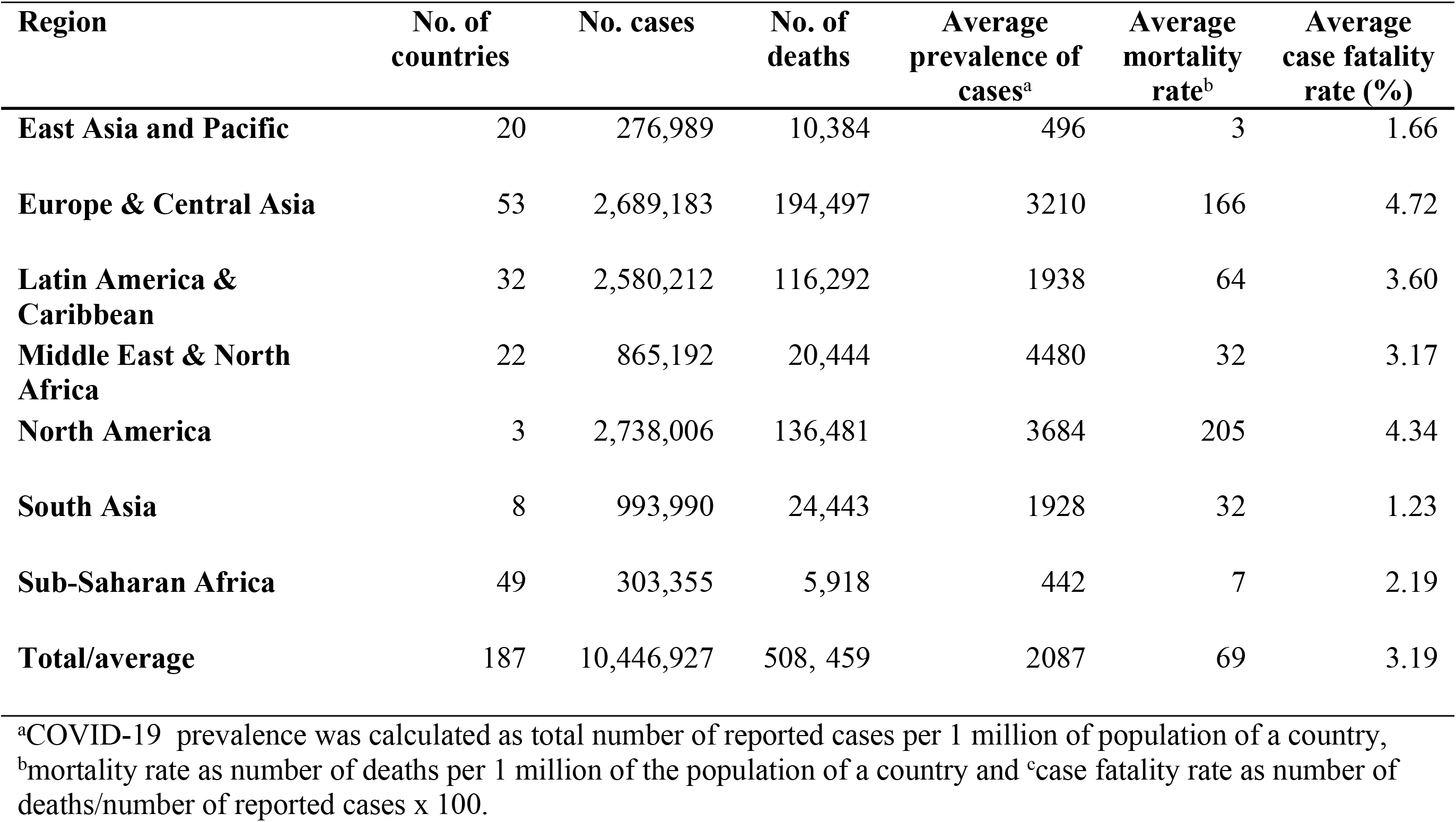
Regional COVID-19 Statistics as of June 30, 2020.

| **Region** | **No. of countries** | **No. cases** | **No. of deaths** | **Average prevalence of cases<sup>a</sup>** | **Average mortality rate<sup>b</sup>** | **Average case fatality rate (%)** |
| --- | --- | --- | --- | --- | --- | --- |
| **East Asia and Pacific** | 20 | 276,989 | 10,384 | 496 | 3 | 1.66 |
| **Europe & Central Asia** | 53 | 2,689,183 | 194,497 | 3210 | 166 | 4.72 |
| **Latin America & Caribbean** | 32 | 2,580,212 | 116,292 | 1938 | 64 | 3.60 |
| **Middle East & North Africa** | 22 | 865,192 | 20,444 | 4480 | 32 | 3.17 |
| **North America** | 3 | 2,738,006 | 136,481 | 3684 | 205 | 4.34 |
| **South Asia** | 8 | 993,990 | 24,443 | 1928 | 32 | 1.23 |
| **Sub-Saharan Africa** | 49 | 303,355 | 5,918 | 442 | 7 | 2.19 |
| **Total/average** | 187 | 10,446,927 | 508, 459 | 2087 | 69 | 3.19 |

<sup>a</sup>COVID-19 prevalence was calculated as total number of reported cases per 1 million of population of a country, <sup>b</sup>mortality rate as number of deaths per 1 million of the population of a country and <sup>c</sup>case fatality rate as number of deaths/number of reported cases x 100.

**Table 2.** Effect of latitude, amount of sunshine and vitamin D status on COVID-19 outcomes.

|  | Latitude |  |  |  | Sunshine |  |  | Vitamin D |  |  |
| --- | --- | --- | --- | --- | --- | --- | --- | --- | --- | --- |
|  | Model | n | Beta (95% CI) | *p* | n | Beta (95% CI) | *p* | n | Beta (95% CI) | *p* |
| Prevalence<sup>a</sup> | Univariate | 182 | 0.022 (0.005, 0.039) | 0.010 | 128 | -2.47 (-3.80, -1.15) | <0.0001 | 74 | -0.018 (-0.041, 0.005) | 0.13 |
|  | Multivariate* | 166 | 0.012 (0.001, 0.024) | 0.041 | 121 | 0.16 (-1.30, 1.62) | 0.83 | 74 | -0.020 (-0.040, 0.001) | 0.060 |
| Mortality rate<sup>b</sup> | Univariate | 182 | 0.056 (0.029, 0.082) | <0.0001 | 128 | -4.44 (-7.24, -1.63) | <0.0001 | 74 | -0.073 (-0.118, -0.028) | 0.002 |
|  | Multivariate* | 166 | 0.042 (0.014, 0.070) | 0.004 | 121 | 0.22 (-3.06, 3.50) | 0.89 | 74 | -0.073 (-0.118, -0.028) | 0.002 |
| Case fatality rate (%)<sup>c</sup> | Univariate | 185 | 0.031 (0.013, 0.050) | 0.001 | 130 | -3.66 (-5.98, -1.34) | 0.0023 | 75 | -0.056 (-0.089, -0.024) | 0.001 |
|  | Multivariate* | 166 | 0.030 (0.013, 0.050) | 0.008 | 122 | -1.51 (-4.44, 1.41) | 0.31 | 74 | -0.054 (-0.089, -0.019) | 0.003 |

n; number of countries included in the analyses <sup>a</sup>COVID-19 prevalence as total number of reported cases per 1 million of population, <sup>b</sup>COVID-19 mortality rate as number of deaths per 1 million of the population and <sup>c</sup>COVID-19 case fatality rate as number of deaths/number of reported cases x 100. \*Multivariate model was adjusted for gross domestic product (GDP), elderly dependency ratio (EDR) and population density.

**Figure 2.**
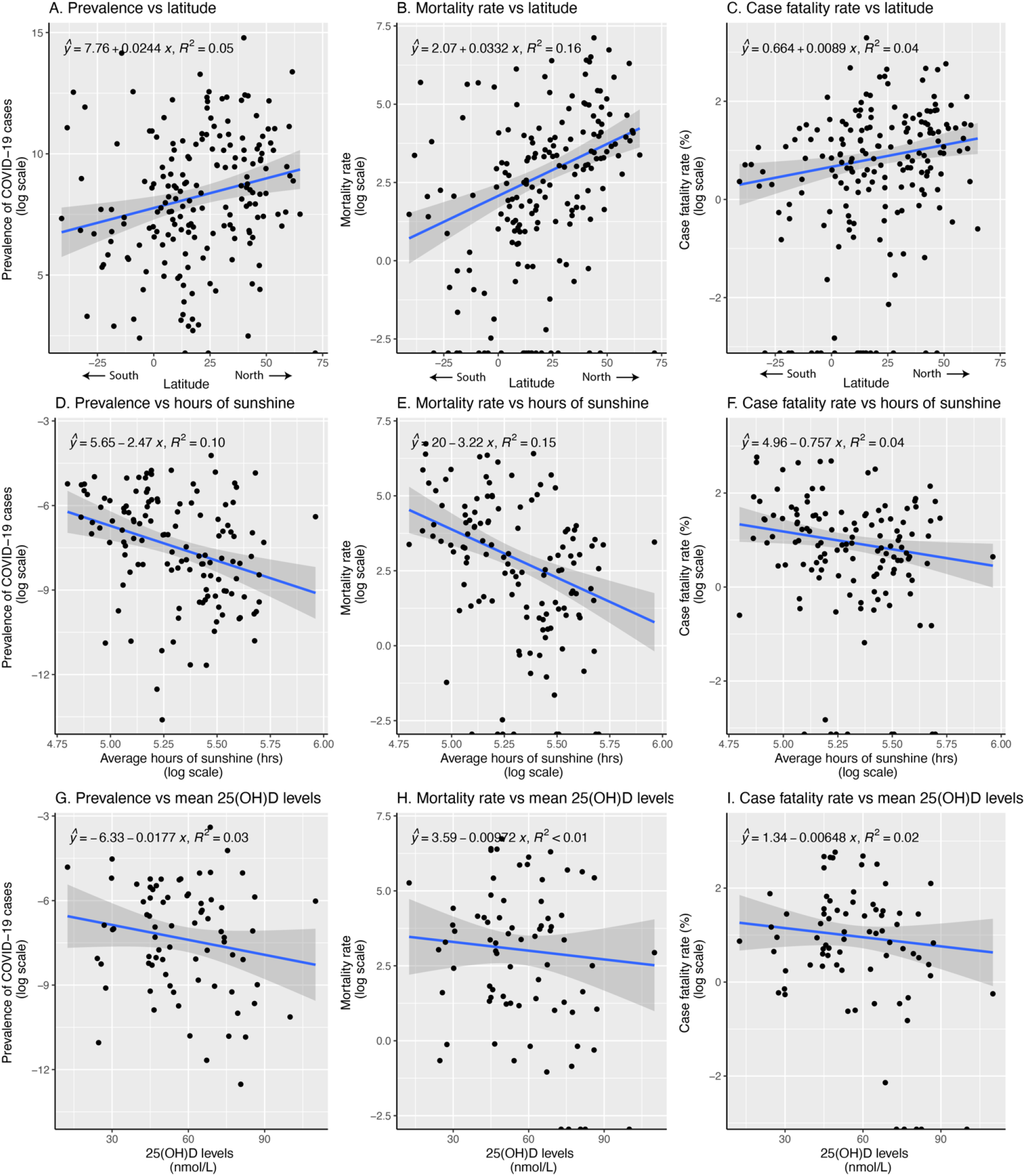
Correlations between COVID-19 metrics and environmental factors across countries. Each point on the plots represents data for an individual country. The COVID-19 prevalence is defined as the total number of reported cases per 1 million population, the mortality rate as the number of deaths per 1 million population, and the case fatality rate as the percentage of reported cases resulting in death, multiplied by 100.

### Effect of sunshine duration on COVID-19 prevalence, mortality rate, and case fatality rate

An inverse association was observed between the average number of sunshine hours and the parameters in univariate analyses but was not significant in multivariate analyses: COVID-19 prevalence rate (β = 0.16; 95% CI: -1.30, 1.62), mortality rate (β = 0.22; 95% CI: -3.06, 3.50), and CFR (β = -1.51; 95% CI: -4.44, 1.41). Similar effects were noted up to the end of March for prevalence and mortality rates, but the association with CFR was not significant. By the end of September, none of these associations remained significant. Scatter plots also indicated an inverse relationship between sunshine duration and COVID-19 metrics (Figure 2D, E and F). Sunshine duration explained 9.9%, 12%, and 1.4% of the variation in COVID-19 prevalence, mortality rate, and CFR, respectively (Figure 2).

### Effect of vitamin D Status on COVID-19 prevalence, mortality rate, and case fatality rate

Mean serum 25OHD) levels were inversely associated with COVID-19 mortality rate (β = – 0.052; 95% CI: –0.094, –0.009) and CFR (β = –0.033; 95% CI: –0.061, –0.006). However, the association between 25OHD levels and COVID-19 prevalence rate was not statistically significant (β = –0.018; 95% CI: –0.041, 0.005; p = 0.13). In March, mean 25OHD levels were significantly associated with mortality rate but not with prevalence or CFR (Supplementary Table 1). By the end of September, significant associations were found between mean 25OHD levels and COVID-19 mortality rate and CFR, but not with prevalence (Supplementary Table 1). Mean 25OHD levels explained 3.2%, 2%, and 2.1% of the variation in COVID-19 prevalence, mortality rate, and CFR, respectively (Figure 2).

## Discussion

This study investigated the associations between geographical factors, vitamin D status, and COVID-19 outcomes across 187 countries during the first six months of 2020. The findings indicate that the prevalence, mortality rate, and case fatality rate (CFR) of COVID-19 were positively associated with latitude and inversely associated with sunshine duration and mean 25OHD levels. These associations were significant up to the end of March 2020 but diminished by the end of September, except for the inverse relationship between mean 25OHD levels and COVID-19 prevalence and CFR. These results suggest potential environmental and nutritional influences on COVID-19 outcomes and highlight a temporal dimension to these relationships.

The positive association between latitude and COVID-19 metrics aligns with previous studies. Rhodes et al. (2020) reported higher mortality rates in countries above 35º North latitude compared to those below, suggesting that latitude may play a role in disease severity^8^. Similarly, higher COVID-19 incidence and transmission rates have been observed in cities and countries farther from the equator^25,26^. Benedetti et al. (2020) found that COVID-19 mortality rates were positively correlated with latitude and negatively correlated with average temperature across regions in the United States and Europe ^27^. The influence of latitude on COVID-19 outcomes may be attributed to lower sunlight exposure during winter months at higher latitudes, leading to vitamin D deficiency ^15^. Additionally, lower temperatures associated with higher latitudes may facilitate viral transmission, as seen in past epidemics like SARS and influenza ^28,29^.

The inverse associations between COVID-19 prevalence, mortality rate, and CFR with average sunshine duration are consistent with previous research. Byass (2020) reported that the incidence of COVID-19 decreased with increasing solar radiation in various locations across China during January and February 2020 ^30^. Furthermore, a study conducted in Jakarta, Indonesia, demonstrated that increased sunlight exposure improved recovery rates among COVID-19 patients^31^. Reduced sunlight exposure may influence COVID-19 outcomes by affecting vitamin D synthesis, which plays a crucial role in immune function^15,16^.

The findings also indicated that mean 25OHD levels were inversely associated with COVID-19 mortality rate and CFR. This observation aligns with Laird et al. (2020), who reported that mean 25OHD levels in European countries were inversely associated with COVID-19 incidence and mortality rates^20^. Additionally, D’Avolio et al. (2020) found that patients who tested positive for SARS-CoV-2 had significantly lower mean 25OHD levels compared to those who tested negative ^32^. Vitamin D deficiency may increase susceptibility to COVID-19 by modulating the immune response and influencing the expression of angiotensin-converting enzyme 2 (ACE2), which is involved in viral entry into cells Munshi^11^. Elevated ACE2 expression has been associated with increased survival in COVID-19 patients^33^. Moreover, vitamin D deficiency is linked to comorbidities such as hypertension, diabetes, and obesity, which are known risk factors for severe COVID-19 outcomes Sattar ^6,34^.

The waning of the observed associations by the end of September may be attributed to the implementation of control measures, changes in testing rates, or seasonal variations. Increased public health interventions, such as lockdowns and social distancing, could have mitigated the influence of environmental factors on COVID-19 transmission ^3,4^. Seasonal changes leading to increased sunlight exposure during the summer months may have improved vitamin D status in populations, potentially affecting disease outcomes Lips ^35,36^.

It is important to acknowledge that, due to the observational nature of this study, causality cannot be inferred, and confounding factors may influence the relationships observed. Other variables such as government policies, healthcare capacity, socioeconomic status, and population behaviors were not accounted for and could confound the results ^8,37^. For instance, Lawal (2021) discussed the paradox of low COVID-19 mortality rates in Africa despite limited healthcare resources, suggesting that factors like younger population demographics and prior exposure to infectious diseases may play a role^8^.

A major strength of this study is the inclusion of data from all countries reporting COVID-19 cases and deaths during the study period, which reduces potential selection bias. Additionally, the analysis considered multiple geographical and environmental factors, including latitude, sunshine duration, and vitamin D status, providing a comprehensive overview of potential influences on COVID-19 outcomes. However, several limitations should be acknowledged. First, the observational design limits the ability to establish causal relationships. Second, sunshine duration and mean 25OHD levels were not available for several countries, which may affect the generalizability of the findings. Third, other potential confounding factors, such as variations in testing rates, reporting practices, public health interventions, and genetic differences among populations, were not accounted for in the analysis. Furthermore, vitamin D levels were based on mean values from existing literature, which may not accurately reflect current population statuses.

This global analysis suggests that higher latitude is associated with increased COVID-19 prevalence, mortality rate, and case fatality rate during the early phase of the pandemic, while higher mean 25-hydroxyvitamin D (25OHD) levels are associated with reduced COVID-19 outcomes. These findings indicate potential environmental and nutritional influences on COVID-19 incidence and disease severity, highlighting vitamin D status as a possible contributing factor. However, recent randomized controlled trials have yielded mixed results regarding the efficacy of vitamin D supplementation in preventing or treating COVID-19.

Some studies have shown no significant benefit of high-dose vitamin D supplementation on hospital length of stay or mortality^38,39^, while others have suggested a reduction in severe outcomes with vitamin D administration^40,41^. Therefore, while observational data support a potential association between vitamin D status and COVID-19 outcomes, causality cannot be established based on current evidence. Considering the ongoing circulation of COVID-19 and the potential for future SARS-like epidemics, understanding the factors that influence disease transmission and severity remains critical. Insights from this study may inform public health strategies aimed at improving population health through safe sun exposure and addressing vitamin D deficiency where prevalent.

## Data Availability

The dataset and accompanying analysis code from the current study are available in our GitHub repository at https://github.com/rmogire/COVID19-Latitude-Sunshine-VitaminD. The dataset is licensed under the Creative Commons Attribution 4.0 International (CC BY 4.0) License, allowing unrestricted use, distribution, and reproduction in any medium, provided the original author and source are credited. The analysis scripts are available under the MIT License, which permits reuse with proper attribution. Researchers and other interested parties are encouraged to access the repository to review the data and replicate the analyses. Any updates or corrections to the dataset are noted in the repository’s changelog. Should there be any questions regarding the data, the corresponding author can be contacted for more information.

## Ethics Statement

This study utilized publicly available, aggregated data at the country level from reputable sources such as the World Health Organization and the World Bank. No individual-level or personally identifiable information was collected or analyzed. Therefore, ethical approval and informed consent were not required.

## Supporting Information

Supplementary Table 1. Effect of latitude, amount of sunshine and vitamin D status on COVID-19 prevalence, mortality rate and case fatality rate by March 31^st^ and September 30^th^

Supplementary Figure 1. Prevalence (number of COVID-19 cases per one million population) of COVID-19 by country as of June 30^th^ 2020.

